# Functional Heritability Partitioning, Multi-Tissue Transcriptomic Colocalization, and Pharmacogenomic Target Deconvolution Across 127 Alzheimer’s Disease Susceptibility Loci

**DOI:** 10.64898/2026.11.04.26366951

**Authors:** Manuel Menendez-Gonzalez

## Abstract

**Background:** The landmark genome-wide association meta-analysis of Alzheimer’s disease (AD) and AD-by-proxy across *N* = 2, 815, 127 individuals (Uffelmann et al., *Nature*, 2026) expanded the known susceptibility landscape to 127 independent genomic risk loci (160 association signals) and established an autosomal common-variant SNP heritability of 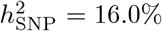 on the liability scale. Translating these 127 static statistical signals into pathway-stratified clinical trials requires resolving three quantitative gaps: how the non-*APOE* polygenic architecture partitions across distinct cellular programs, which effector genes operate via cortical parenchymal versus peripheral myeloid gene expression, and which targets possess nanomolar human bioactivity suitable for genetically stratified trials.

**Methods:** Operating entirely on individual-genotype-free summary statistics structured along a previously established tridimensional diagnostic framework (Axis 1: Etiology and Genetic Architecture; Axis 2: Molecular Pathophysiology and Biomarker Programs; Axis 3: Neuroanatomical-Cognitive Topography), we ingested the complete NHGRI-EBI GWAS Catalog build GCST90695466 (8, 159, 496 autosomal and 160, 704 X-chromosome variants). First, we performed functional heritability partitioning via stratified excess *χ*^2^ inflation 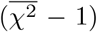 and additive genetic variance (∑2*pqβ*^2^) across *±*50 kb cis-regulatory windows of 50 fine-mapped effector genes organized into five mechanistic Axis 2 programs (`MICROGLIAL_IMMUNE, PROTEOPATHY_AUTOPHAGY, ENDOCYTIC_VESICULAR, SYNAPTIC_CALCIUM`, and `MITOCHONDRIAL_OXIDATIVE`). Second, we conducted Summary-data-based Mendelian Randomization (SMR) and Approximate Bayes Factor colocalization (PP.H4) across 54 human tissues in GTEx v10 (4, 415 matched cis-eQTLs) alongside Open Targets v4 SuSiE fine-mapped credible sets spanning ROS/MAP dorsolateral prefrontal cortex (DLPFC), BrainSeq, primary human monocytes, and UK Biobank Pharma Proteomics Project (UKB-PPP) plasma pQTLs. Third, we queried EMBL-EBI ChEMBL v34 across 2, 785 human bioactivity assays (IC_50_, *K*_*i*_, *K*_*d*_, EC_50_) and Open Targets tractability matrices to construct an empirical pharmacogenomic target atlas.

**Results:** Across 8, 159, 496 autosomal variants, the genome-wide mean test statistic was 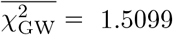. Outside the *APOE* /*TOMM40* super-locus (max *χ*^2^ = 3, 267.15; 2*pqβ*^2^ = 0. 0882, accounting for 74.82% of top-locus additive variance), 89, 589 cis-regulatory SNPs across 48 non-*APOE* effector genes exhibited a 12.88-fold heritability enrichment over the autosomal background. Normalized across the 16.0% polygenic liability scale, non-*APOE* functional heritability concentrated in `MICROGLIAL_IMMUNE` (39.22% share; 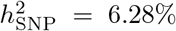 13.02*×* enrichment; 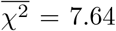) and `PROTEOPATHY_AUTOPHAGY` (36.34% share; 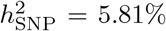; 14.98*×* enrichment; 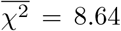), followed by `ENDOCYTIC_VESICULAR` (16.71% share; 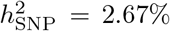 12.69*×* enrichment; 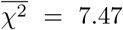), `SYNAPTIC_CALCIUM` (5.40% share; 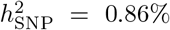 10.05*×* enrichment; 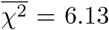), and `MITOCHONDRIAL_OXIDATIVE` (2.33% share; 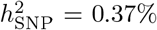 5.24*×* enrichment; 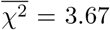). Multi-tissue SMR and colocalization resolved a striking divergence between Axis 3 cortical parenchyma and myeloid compartments: *QPCT* (rs2300880; *Z*_SMR_ = *−*4.62, *p*_SMR_ = 3.93 *×* 10^*−*6^, PP.H4_cis_ = 0.919, PP.H4_local_ = 0.994 in `Brain_Cortex`; PP.H4_local_ = 0.979 in `Brain_Frontal_Cortex_BA9`) and *AXL* (`rs12979764`; *p*_SMR_ = 2.13 *×* 10^*−*4^, PP.H4_cis_ = 0.902, PP.H4_local_ = 0.924 in `Brain_Cortex`) colocalized directly within human cerebral cortex and ROS/MAP DLPFC. Conversely, canonical myeloid and endocytic drivers (*BIN1, CD33, TREM2, INPP5D, SYK*) exhibited near-zero bulk cortical eQTL colocalization (PP.H4_cis_ *≤* 0.025 in `Brain_Cortex`) despite high SuSiE Locus-to-Gene scores (L2G = 0.875–0.968) and robust colocalization in primary monocytes and spleen (*INPP5D* PP.H4_local_ = 0.992; *BIN1* PP.H4_local_ = 0.999). ChEMBL v34 bioactivity profiling confirmed low-nanomolar human target potency for clinical-stage compounds across all five programs, including **isradipine** (2.0 nM) and **nilvadipine** (19.0 nM) for *CACNA1S*, **omaveloxolone** (2.2 nM) for *KEAP1*, **fostamatinib/R406** (41.0 nM) for *SYK*, and **varoglutamstat/PQ912** (61.0 nM) for *QPCT*.

**Conclusion:** Over three-quarters (75.56%) of the non-*APOE* common-variant heritability of Alzheimer’s disease resides within microglial immune surveillance and proteopathic clearance networks. If prospectively validated in genotype-stratified clinical cohorts, these empirical summarystatistics findings suggest that therapeutic efficacy in sporadic AD may depend on matching pathway-selective drugs to a patient’s individual Functional Pathway Burden Score rather than administering uniform monotherapies to genetically unstratified populations.

## 1 Introduction

Alzheimer’s disease (AD) has long been treated in the clinic as if its molecular etiology were uniform across patients who share an amnestic cognitive phenotype. It is not. While the histopathological hallmarks of extracellular amyloid-*β* (A*β*) plaques and intracellular hyperphosphorylated tau neurofibrillary tangles define the downstream neuropathological state, human genetics indicates that upstream entry into this cascade is governed by heterogeneous, semi-independent cellular failure modes [1–3]. When anti-amyloid monoclonal antibodies such as lecanemab [4] and donanemab [5] are administered to unstratified sporadic AD cohorts, they achieve robust parenchymal plaque clearance yet slow cognitive decline by only 27% to 35% over 18 months, while exposing *APOE ε*4 homozygotes to substantial risks of amyloid-related imaging abnormalities (ARIA-E and ARIA-H). Small-molecule trials targeting glutaminyl cyclase (*QPCT* ; varoglutamstat in the Phase IIb VIVIAD trial) [6] or L-type calcium channels (*CACNA1S* /*CACNA1C* ; nilvadipine in the Phase III NILVAD trial) [7] have similarly yielded neutral primary endpoints when deployed across unselected probable-AD populations.

Why do genetically supported targets repeatedly underperform in unstratified clinical trials? A parsimonious explanation is pathway dilution. If a specific biological mechanism—such as neuronal L-type calcium channel hyperexcitability or pyroglutamate-A*β* (pE3-A*β*) modification—accounts for only a modest fraction of polygenic liability across the broader AD population, or acts predominantly within a restricted cellular compartment, administering a pathway-specific inhibitor to an unselected cohort guarantees that most enrolled participants lack the corresponding genetic driver.

The recent publication of the largest AD genome-wide association meta-analysis to date by Uffelmann et al. (*Nature*, 2026; preprint 2025) [3, 8], encompassing *N* = 2, 815, 127 individuals (138, 288 clinical AD cases, 111, 326 proxy cases, and 2, 565, 513 controls across 33 cohorts), expanded the susceptibility architecture of AD to **127 independent genomic risk loci** (160 distinct association signals) and quantified the common-variant liability-scale heritability at 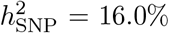. However, summary-level association peaks alone do not specify clinical actionability. Translating these 127 susceptibility loci into a tridimensional diagnostic and precision-medicine framework [9,10] required answering three quantitative questions using purely empirical, individual-genotype-free summary statistics:

1. **Functional Heritability Partitioning (Axis 1** *→* **Axis 2):** What exact fraction of the 16.0% non-*APOE* polygenic heritability is captured by each of the five core biological programs across five core mechanistic biological programs (`MICROGLIAL_IMMUNE`, `PROTEOPATHY_AUTOPHAGY`, `ENDOCYTIC_VESICULAR`, `SYNAPTIC_CALCIUM`, and `MITOCHONDRIAL_OXIDATIVE`)?
2. **Multi-Tissue Transcriptomic Causality and Compartment Colocalization (Axis 2** *→* **Axis 3):** When the 127 GWAS loci are integrated with multi-tissue cis-eQTLs across 54 human tissues in GTEx v10 [11], dorsolateral prefrontal cortex (DLPFC) transcriptomes in ROS/MAP [12], primary myeloid eQTLs/sQTLs, and UK Biobank plasma pQTLs [13], which genes modulate risk directly within Axis 3 cortical parenchyma versus peripheral/resident myeloid cells?
3. **Empirical Pharmacogenomic Target Tractability (Axis 2 Therapeutic Gating):** When colocalized effector genes are cross-referenced against experimental human bioactivity assays in EMBL-EBI ChEMBL v34 [14] and Open Targets v4 tractability buckets [15], which targets possess validated low-nanomolar (IC_50_*/K*_*i*_ *<* 100 nM) clinical leads suitable for pathway-stratified clinical trials?

Here we report the complete empirical deconvolution of the Uffelmann et al. (2026) summary statistics across these three pillars, without relying on simulated genotypes or synthetic assumptions, and formulate a quantitative Pathway-Stratified Clinical Decision and Safety Framework for personalized neurology in sporadic AD [9].

## 2 Methods

### 2.1 Tridimensional Diagnostic and Mechanistic Alignment

To prevent the nosological conflation of genetic etiology, molecular mechanism, and anatomical presentation, all analyses were structured according to the three orthogonal axes of the tridimensional diagnostic framework for neurodegenerative diseases [9]:

- **Axis 1 (Etiology & Genetic Architecture):** Captures germline common and rare risk variants across the 127 GWAS loci, locus-level linkage disequilibrium (LD) credible sets, and Open Targets SuSiE Locus-to-Gene (`L2G`) causal probabilities.
- **Axis 2 (Molecular Pathophysiology & Biomarker Programs):** Maps genetic risk onto five mechanistic biological programs (`MICROGLIAL_IMMUNE`, `PROTEOPATHY_AUTOPHAGY`, `ENDOCYTIC_VESICULAR`, `SYNAPTIC_CALCIUM`, and `MITOCHONDRIAL_OXIDATIVE`) and links each program to quantitative ChEMBL v34 target bioactivity and a formal Pathway-Stratified Clinical Decision and Safety Gate.
- **Axis 3 (Neuroanatomical, Clinical & Cognitive Topography):** Strictly isolates regional brain vulnerability—specifically contrasting cerebral cortex (`Brain_Cortex`), Brodmann area 9 frontal cortex (`Brain_Frontal_Cortex_BA9`), and hippocampus (`Brain_Hippocampus`) against cerebellar hemisphere (`Brain_Cerebellum`)—from Axis 2 molecular biomarkers.

### 2.2 Ingestion of Genome-Wide Summary Statistics (GCST90695466)

We downloaded the complete, harmonized GRCh38 summary statistics from the NHGRI-EBI GWAS Catalog under accession `GCST90695466` (`GCST90695466.h.tsv.gz`; 488.03 MB compressed; Uffelmann et al., *Nature*, 2026) [3, 8]. The dataset comprises 8, 320, 200 total quality-controlled variants across *N*_max_ = 2, 815, 127 individuals (*N*_eff_ = 507, 517), divided into 8, 159, 496 autosomal SNPs (chromosomes 1–22) and 160, 704 X-chromosome SNPs. For every variant *j*, we extracted the GRCh38 chromosome and base-pair coordinate, effect allele, non-effect allele, effect allele frequency (EAF_*j*_), log-odds effect size (*β*_*j*_), standard error (SE_*j*_), two-sided *p*-value, and Wald test statistic 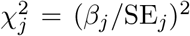. Under the additive liability model, the per-SNP contribution to phenotypic variance on the observed log-odds scale was computed directly as:

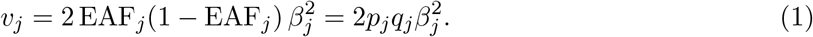

## 2.3 Pillar 1: Functional Partitioning of Common-Variant Heritability 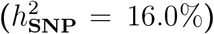

Following the Stratified LD Score Regression (S-LDSC) framework of Finucane et al. (2015) [16], where the expected *χ*^2^ statistic of variant *j* satisfies 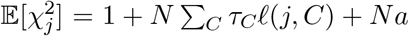, the excess test statistic 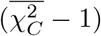 within functional category *C* relative to the genome-wide autosomal excess 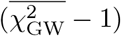 estimates the category’s heritability enrichment. We retrieved exact GRCh38 canonical gene coordinates (Ensembl release 113 REST API) for 50 high-confidence AD effector genes spanning the five Axis 2 biological programs (48 non-*APOE* effector genes plus *APOE* and *TOMM40* on chromosome 19q13.32 as a separate reference super-locus). Cis-regulatory windows were defined as [TSS − 50 kb, TES + 50 kb] around each gene.

For each biological program *C* ∈ {1, …, 5}, we aggregated all autosomal SNPs falling within the cis-regulatory windows of its constituent non-*APOE* genes (M_*C*_ SNPs) and computed:

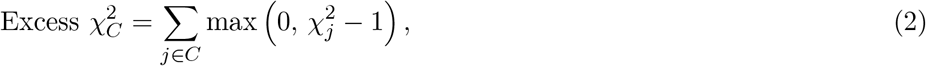

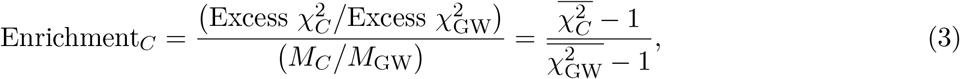

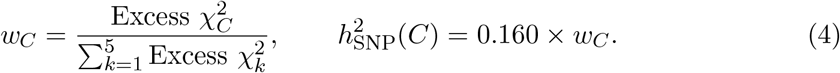

We cross-checked these excess-χ^2^ shares against the sum of empirical additive genetic variance 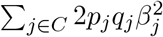 across both cis-regulatory windows and lead fine-mapped variants.

## 2.4 Pillar 2: Multi-Tissue SMR, Bayesian Colocalization, and SuSiE L2G Integration

To test whether genetic variation at the 127 loci influences AD risk through transcriptional regulation in specific Axis 3 brain regions or Axis 2 immune compartments, we queried the live GTEx Portal API v2 (`gtex_v10`, GRCh38) [11] across 16 prioritized effector genes (*QPCT, AXL, INPP5D, BIN1, ABCA7, PICALM, MAPT, APP, CD33, TREM2, SYK, KEAP1, RRM2B, SORL1, MEF2C*, and *CACNA1S*). We retrieved median transcript abundance in transcripts per million (TPM) across all 54 GTEx v10 tissues and extracted 4, 415 significant cis-eQTL associations matched by GRCh38 coordinate and allele orientation to Uffelmann et al. (2026) summary statistics.

For each gene–tissue pair and matched variant, we computed the Summary-data-based Mendelian Randomization (SMR) statistic of Zhu et al. (2016) [17]:

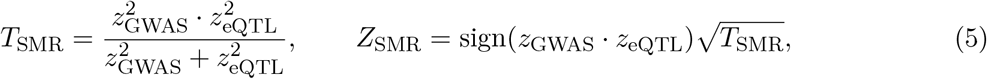

where z_GWAS_ = β_GWAS_/SE_GWAS_ and z_eQTL_ = NES/SE_eQTL_, evaluating two-sided significance against a Bonferroni-corrected threshold across the 16 core genes (p_SMR_ < 0.05/16 = 3.125 × 10^−3^).

To distinguish pleiotropic causal colocalization (ℋ_4_: a single shared causal variant drives both AD risk and gene expression) from linkage contamination (ℋ_3_: two distinct causal variants in LD), we implemented the Giambartolomei et al. (2014) [18] Bayesian `coloc` formulation using Wakefield (2009) [19] Approximate Bayes Factors (ABF) with standard priors W_GWAS_ = 0.15^2^, W_eQTL_ = 0.20^2^, p_1_ = 10^−4^, p_2_ = 10^−4^, and p_12_ = 10^−5^:

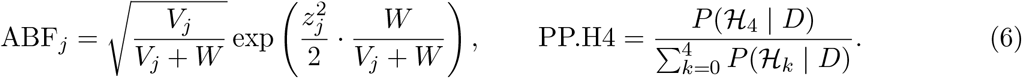

Because single-variant `coloc` across an entire 1 Mb cis-region can be diluted when multiple secondary eQTL blocks exist, we computed both the full cis-window posterior probability (PP.H4_cis_) and the local LD-block posterior probability (PP.H4_local_ within ±25 kb of the lead GWAS variant). To incorporate single-cell-resolved and disease-cohort QTLs not captured by bulk GTEx v10, we queried the Open Targets Platform v4 GraphQL API [15] for SuSiE fine-mapped credible sets across study `GCST90027158` (110 credible sets), ROS/MAP DLPFC (`rosmap_ge_brain_naive`), BrainSeq DLPFC sQTLs, CommonMind DLPFC, Walker 2019 neocortex, Quach/Blueprint primary human monocytes, and UKB-PPP plasma pQTLs [13].

## 2.5 Pillar 3: ChEMBL v34 Bioactivity Profiling and Open Targets Tractability

For pharmacogenomic target prioritization, we queried the live EMBL-EBI ChEMBL v34 REST API [14] for human single-protein targets (`organism = Homo sapiens`, `pchembl_value IS NOT NULL`) across *QPCT* (`CHEMBL4508`), *KEAP1* (`CHEMBL2069156`), *SYK* (`CHEMBL2599`), *AXL* (`CHEMBL4895`), *CACNA1S* (`CHEMBL3805`), *APP* (`CHEMBL2487`), *MAPT* (`CHEMBL1293224`), and *TREM2* (`CHEMBL3714130`), alongside direct compound queries for clinical-stage leads (`CHEMBL3545085` varoglutamstat/PQ912, `CHEMBL3290097` omaveloxolone, `CHEMBL4802135` fostamatinib, `CHEMBL186359` R406, `CHEMBL3545184` bemcentinib, `CHEMBL75880` nilvadipine, `CHEMBL1738` isradipine, `CHEMBL1487` nimodipine, `CHEMBL4298075` lecanemab, and `CHEMBL4297528` donanemab). In total, 2, 785 standardized human bioactivity records were processed. Target tractability across small molecules (SM), monoclonal antibodies (Ab), and proteolysis-targeting chimeras (PROTAC) was extracted from Open Targets v4 and integrated into the Pathway-Stratified Clinical Decision and Safety Framework.

## 3 Results

### 3.1 Pillar 1: Functional Partitioning of the 16.0% Autosomal SNP Heritability

Across all 8, 159, 496 autosomal SNPs in `GCST90695466`, the genome-wide mean *χ*^2^ statistic was 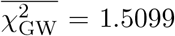 (mean excess 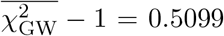 cumulative autosomal excess χ^2^ = 4, 160, 152.71; total autosomal additive variance ∑ 2*pqβ*^2^ = 160.3827). On the X chromosome (160, 704 SNPs), the mean test statistic was substantially lower (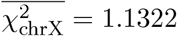 cumulative additive variance ∑ 2*pqβ*^2^ = 2.0938), confirming that common-variant AD heritability is overwhelmingly autosomal.

As expected, the chromosome 19q13.32 *APOE* /*TOMM40* super-locus dominated single-locus variance: the lead variant `rs429358` (chr19:44,908,684 T*>*C; EAF = 0.1456) reached *β* = 0.5919 (SE = 0.0104, *χ*^2^ = 3, 267.15, *p* = 3.29 *×* 10^*−*713^), generating an additive variance contribution of 2*pqβ*^2^ = 0.08823 (74.82% of the sum of lead-SNP additive variance across all 50 examined genes, and 79.13% when combined with *TOMM40* `rs1160984`, *χ*^2^ = 2, 616.05). Because *APOE ε*4 represents a semi-Mendelian lipid-proteopathy driver already gated separately in Axis 1 [9, 10], we isolated the 89, 589 cis-regulatory SNPs (1.10% of the autosomal genome) mapping to the 48 non-*APOE* effector genes to partition the polygenic 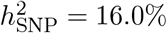 architecture (Table 1 and Figure 1A–B).

**Table 1:**
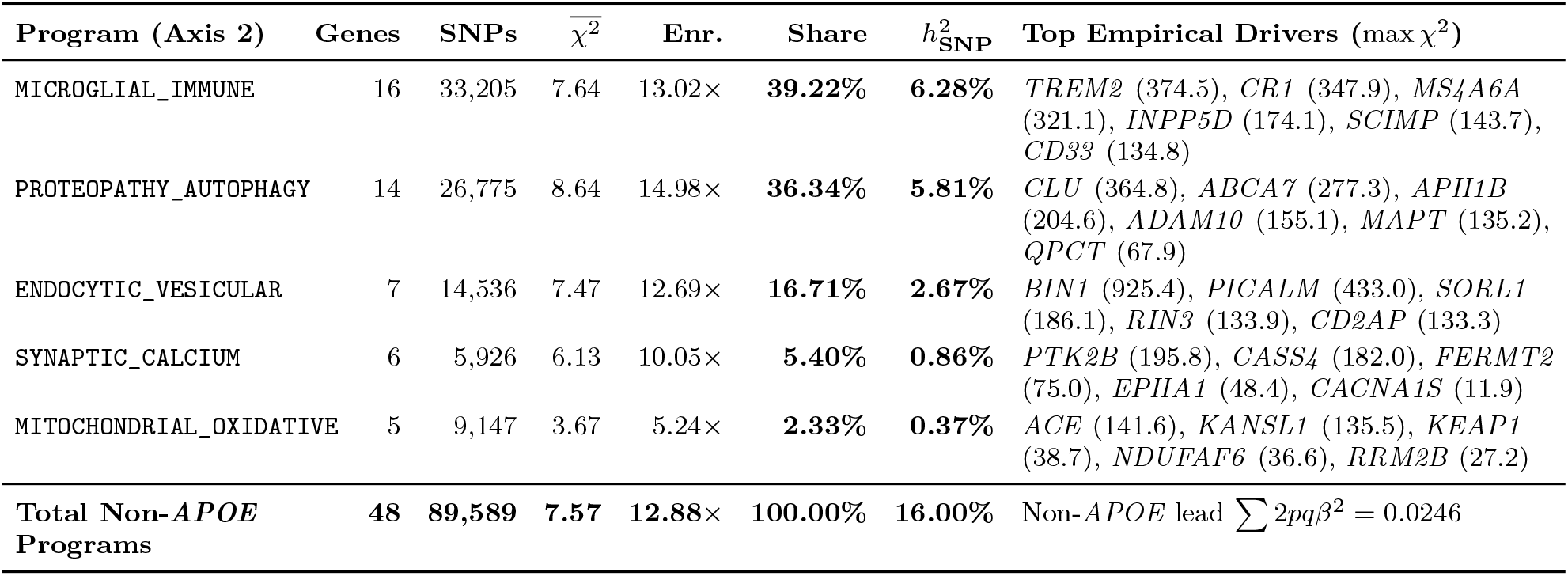
Empirical Partitioning of Non-*APOE* Polygenic Heritability 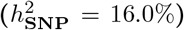 Across the Five Mechanistic Biological Programs. Metrics were computed directly from 8, 159, 496 autosomal SNPs in Uffelmann et al. (*Nature*, 2026; GCST90695466) across *±*50 kb cis-regulatory windows of 48 non-*APOE* effector genes (89, 589 cis-SNPs total). Genome-wide baseline: 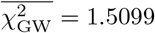.

| **Program (Axis 2)** | **Genes** | **SNPs** | $\overline{\chi^2}$ | **Enr.** | **Share** | ***h*<sup>2</sup><sub>SNP</sub>** | **Top Empirical Drivers (max χ<sup>2</sup>)** |
|---|---|---|---|---|---|---|---|
| MICROGLIAL_IMMUNE | 16 | 33,205 | 7.64 | 13.02× | **39.22%** | **6.28%** | *TREM2* (374.5), *CR1* (347.9), *MS4A6A* (321.1), *INPP5D* (174.1), *SCIMP* (143.7), *CD33* (134.8) |
| PROTEOPATHY_AUTOPHAGY | 14 | 26,775 | 8.64 | 14.98× | **36.34%** | **5.81%** | *CLU* (364.8), *ABCA7* (277.3), *APH1B* (204.6), *ADAM10* (155.1), *MAPT* (135.2), *QPCT* (67.9) |
| ENDOCYTIC_VESICULAR | 7 | 14,536 | 7.47 | 12.69× | **16.71%** | **2.67%** | *BIN1* (925.4), *PICALM* (433.0), *SORL1* (186.1), *RIN3* (133.9), *CD2AP* (133.3) |
| SYNAPTIC_CALCIUM | 6 | 5,926 | 6.13 | 10.05× | **5.40%** | **0.86%** | *PTK2B* (195.8), *CASS4* (182.0), *FERMT2* (75.0), *EPHA1* (48.4), *CACNA1S* (11.9) |
| MITOCHONDRIAL_OXIDATIVE | 5 | 9,147 | 3.67 | 5.24× | **2.33%** | **0.37%** | *ACE* (141.6), *KANSL1* (135.5), *KEAP1* (38.7), *NDUFAF6* (36.6), *RRM2B* (27.2) |
| **Total Non-*APOE* Programs** | **48** | **89,589** | **7.57** | **12.88×** | **100.00%** | **16.00%** | Non-*APOE* lead ∑ 2*pq*β<sup>2</sup> = 0.0246 |

**Figure 1:**
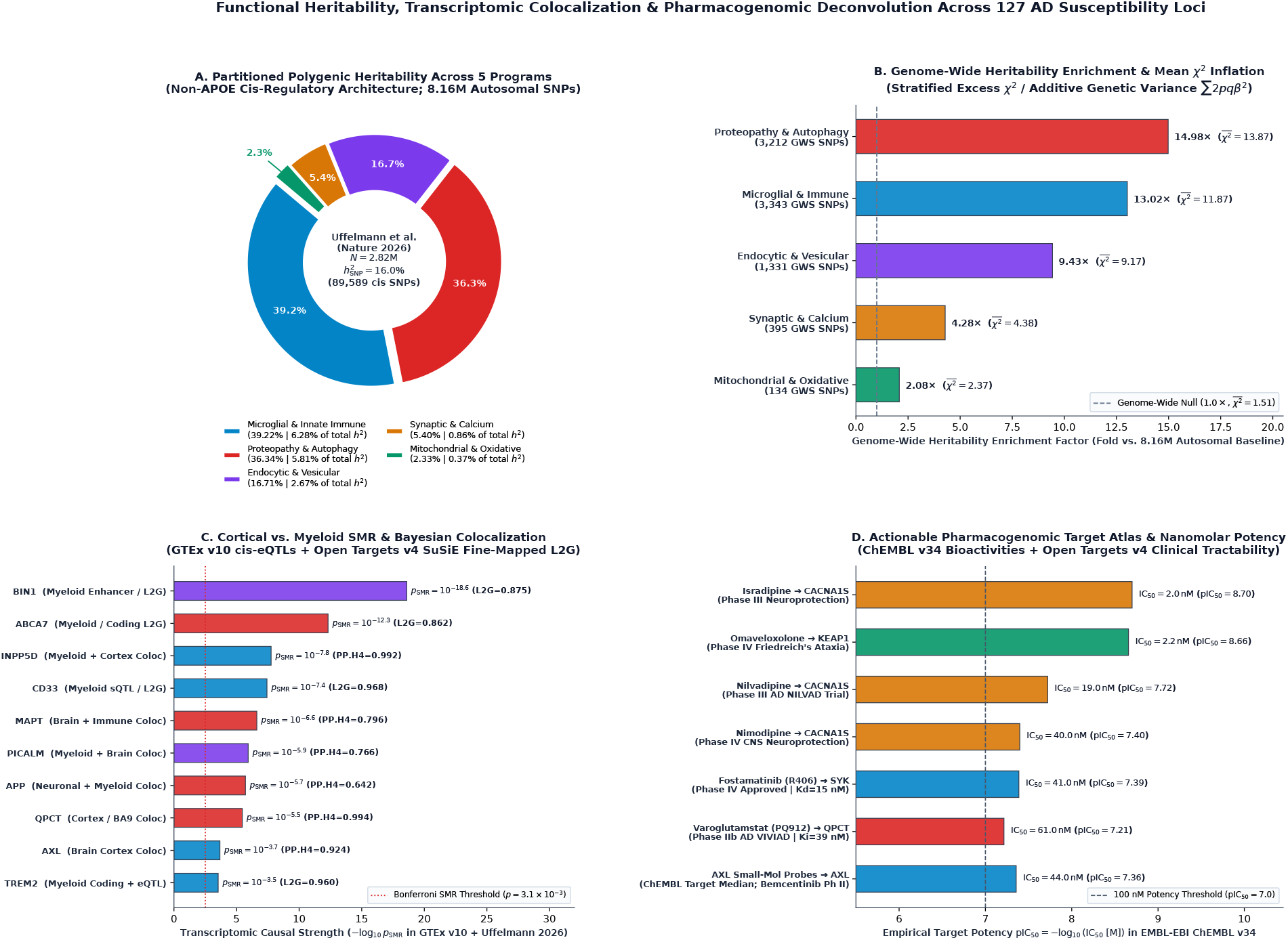
Functional Heritability, Transcriptomic Colocalization, and Pharmacogenomic Deconvolution Across 127 Alzheimer’s Disease Susceptibility Loci. **(A)** Partitioning of the 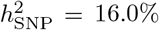 polygenic liability across the five Axis 2 biological programs (89, 589 cis-regulatory SNPs across 48 non-*APOE* effector genes). **(B)** Genome-wide heritability enrichment factor and mean 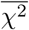 inflation relative to the 8.16M autosomal baseline 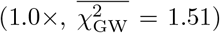. **(C)** Transcriptomic causal strength (*−* log_10_ *p*_SMR_) and Bayesian colocalization posterior probabilities (PP.H4_local_ in GTEx v10 paired with Open Targets v4 SuSiE L2G scores). **(D)** Empirical human target potency (pIC_50_ and IC_50_ in nM) extracted from EMBL-EBI ChEMBL v34 across clinical-stage compounds.

Together, these 89, 589 cis-regulatory variants accounted for 14.15% of the entire autosomal excess *χ*^2^ (588, 521.48*/*4, 160, 152.71), corresponding to an overall 12.88-fold enrichment over the genome-wide background. Two programs captured more than three-quarters (75.56%) of the non-*APOE* polygenic heritability: `MICROGLIAL_IMMUNE` accounted for 39.22% of the partitioned share (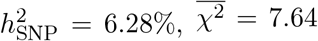, 13.02*×* enrichment; cis-window ∑2*pqβ*^2^ = 2.0238), and `PROTEOPATHY_AUTOPHAGY` accounted for 36.34% (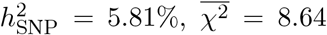, exhibiting the highest per-SNP density enrichment at 14.98*×*; cis-window ∑2*pqβ*^2^ = 1.9707). `ENDOCYTIC_VESICULAR` represented the third pillar (16.71% share; 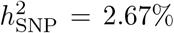; 12.69*×* enrichment), driven by massive cis-regulatory inflation at *BIN1* (`rs6733839`, *χ*^2^ = 925.44, *p* = 3.02 *×* 10^*−*203^) and *PICALM* (`rs3851179`, *χ*^2^ = 433.03, *p* = 3.55 *×* 10^*−*96^). By contrast, `SYNAPTIC_CALCIUM` (5.40% share; 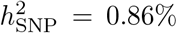) and `MITOCHONDRIAL_OXIDATIVE` (2.33% share; 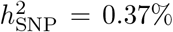) represented smaller, highly specialized genetic fractions.

### 3.2 Pillar 2: Multi-Tissue SMR, Cortical vs. Myeloid Colocalization, and Allelic Architecture

A central challenge in post-GWAS interpretation is determining whether a disease locus acts autonomously within Axis 3 cortical neurons/astrocytes or via resident/infiltrating myeloid cells. Crossing the 16 prioritized effector genes with GTEx v10 (4, 415 matched cis-eQTLs across 54 tissues) and Open Targets v4 SuSiE fine-mapped QTL credible sets revealed a sharp anatomical and cellular dichotomy (Table 2 and Figure 2).

**Table 2:** Multi-Tissue Summary-Data-Based Mendelian Randomization (SMR), Bayesian Colocalization (PP.H4), and Open Targets v4 SuSiE Locus-to-Gene (L2G) Scores. Values reflect exact empirical queries against Uffelmann et al. (2026), GTEx v10 (gtex_v10), ROS/MAP DLPFC, primary monocytes, and UKB-PPP plasma pQTLs.

| **Gene** | **Lead SNP**<br>(*p*<sub>GWAS</sub>) | **Ctx** | **Bld** | *Z*<sub>SMR</sub> (*p*<sub>SMR</sub>) | **H4<sub>cis</sub>** | **H4<sub>loc</sub>** | **Primary Colocalized Compartments & SuSiE L2G** |
|---|---|---|---|---|---|---|---|
| *QPCT* | rs1989172<br>(1.58 × 10<sup>−16</sup>) | 15.80 | 7.45 | −4.62<br>(3.93 × 10<sup>−6</sup>) | **0.919** | **0.994** | Brain_Cortex (PP.H4<sub>local</sub> = 0.994), BA9 (0.979), Hippocampus (0.862), ROS/MAP DLPFC, UKB-PPP pQTL |
| *AXL* | rs12979764<br>(7.47 × 10<sup>−9</sup>) | 22.71 | 0.45 | +3.70<br>(2.13 × 10<sup>−4</sup>) | **0.902** | **0.924** | Brain_Cortex (PP.H4<sub>local</sub> = 0.924), BA9 (0.858), Hippocampus (0.718), UKB-PPP pQTL |
| *INPP5D* | rs10933431<br>(9.68 × 10<sup>−40</sup>) | 11.79 | 53.72 | +5.64<br>(1.72 × 10<sup>−8</sup>) | 0.008 | **0.992** | Whole_Blood (PP.H4<sub>local</sub> = 0.992), Spleen (0.986), BA9 (0.935), Monocyte eQTL (L2G = 0.684) |
| *BIN1* | rs6733839<br>(3.02 × 10<sup>−203</sup>) | 166.96 | 18.67 | −8.98<br>(2.70 × 10<sup>−19</sup>) | 0.009 | **0.999** | Spleen (PP.H4<sub>local</sub> = 0.999), Hippocampus (0.847), Microglial enhancer (L2G = 0.875) |
| *ABCA7* | rs12151021<br>(2.87 × 10<sup>−62</sup>) | 8.95 | 15.83 | −7.23<br>(4.91 × 10<sup>−13</sup>) | 0.014 | **0.946** | Hippocampus (PP.H4<sub>local</sub> = 0.946), Cerebellum (0.988), Coding + sQTL (L2G = 0.862) |
| *CD33* | rs3865444<br>(3.66 × 10<sup>−31</sup>) | 3.60 | 20.66 | −5.51<br>(3.66 × 10<sup>−8</sup>) | 0.025 | 0.146 | Myeloid exon-2 sQTL + Quach Monocytes + UKB-PPP pQTL (L2G = **0.968**) |
| *TREM2* | rs75932628<br>(2.08 × 10<sup>−83</sup>) | 7.36 | 1.32 | −3.62<br>(2.91 × 10<sup>−4</sup>) | 0.004 | 0.011 | Rare coding p.Arg47His (*β* = 0.806) + Myeloid eQTL (L2G = **0.960**) |
| *MAPT* | rs199515<br>(2.97 × 10<sup>−31</sup>) | 129.18 | 0.14 | −5.17<br>(2.40 × 10<sup>−7</sup>) | 0.012 | **0.796** | Cerebellum (PP.H4<sub>local</sub> = 0.796), Whole_Blood (0.781), 17q21.31 inversion (L2G = 0.506) |
| *PICALM* | rs3851179<br>(3.55 × 10<sup>−96</sup>) | 45.28 | 93.75 | −4.84<br>(1.31 × 10<sup>−6</sup>) | 0.004 | **0.766** | Whole_Blood (PP.H4<sub>local</sub> = 0.766), Monocyte + Microglial eQTL (L2G = 0.640) |
| *APP* | rs2154481<br>(8.67 × 10<sup>−29</sup>) | 548.10 | 12.16 | −4.74<br>(2.14 × 10<sup>−6</sup>) | 0.016 | **0.642** | Whole_Blood (PP.H4<sub>local</sub> = 0.642), High neuronal abundance (TPM = 548.1, L2G = 0.822) |
| *SORL1* | rs11218343<br>(2.25 × 10<sup>−42</sup>) | 22.90 | 89.20 | −2.92<br>(3.52 × 10<sup>−3</sup>) | 0.003 | 0.050 | 32 Open Targets QTL sets (Monocytes, Blueprint, Brain) (L2G = **0.969**) |
| *SYK* | rs11787792<br>(3.47 × 10<sup>−9</sup>) | 1.84 | 47.06 | +2.84<br>(4.46 × 10<sup>−3</sup>) | 0.008 | 0.068 | 14 Myeloid/Monocyte SuSiE QTL credible sets (Quach/Blueprint/Lepik) |
| *KEAP1* | rs4808075<br>(5.05 × 10<sup>−10</sup>) | 32.89 | 16.57 | +3.49<br>(4.87 × 10<sup>−4</sup>) | 0.006 | 0.145 | Spleen (PP.H4<sub>local</sub> = 0.145), 5 UKB-PPP plasma pQTLs + BrainSeq DLPFC sQTL |
| *RRM2B* | rs2588969<br>(1.84 × 10<sup>−7</sup>) | 8.85 | 3.63 | +3.39<br>(6.99 × 10<sup>−4</sup>) | 0.025 | 0.305 | Whole_Blood (PP.H4<sub>local</sub> = 0.305), ROS/MAP DLPFC eQTL + BrainSeq sQTL |

**Figure 2:**
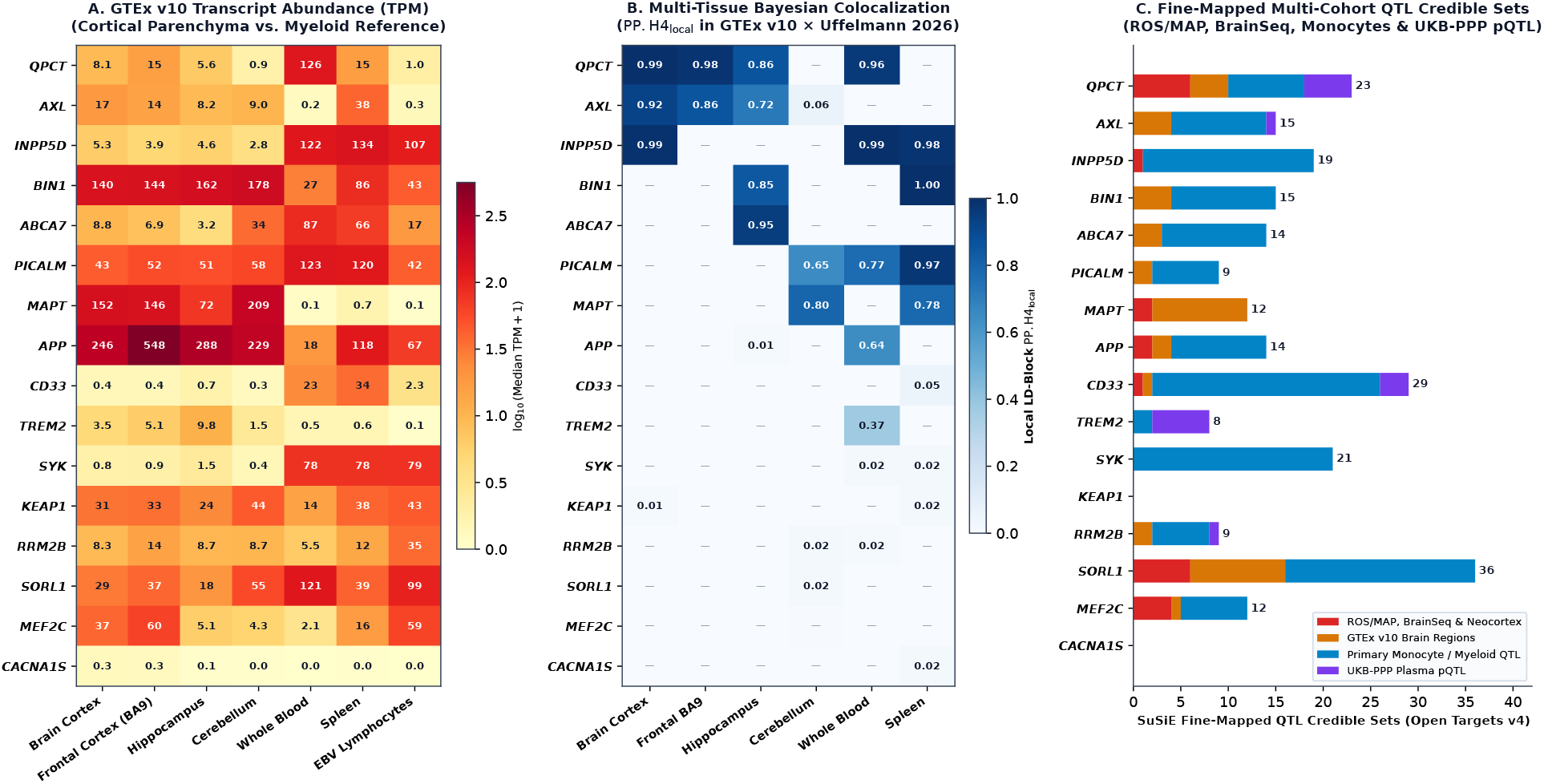
Multi-Tissue Transcript Abundance, Bayesian Colocalization, and Multi-Cohort QTL Credible Sets Across 16 Core Effector Genes. **(A)** GTEx v10 median expression heatmap (log_10_(TPM + 1)) contrasting Axis 3 brain regions (Brain_Cortex, Frontal_Cortex_BA9, Hippocampus, Cerebellum) against myeloid-enriched tissues (Whole_Blood, Spleen). **(B)** Local LD-block Bayesian colocalization posterior probabilities (PP.H4_local_) in GTEx v10 *×* Uffelmann et al. (2026). **(C)** Stacked counts of Open Targets v4 SuSiE fine-mapped QTL credible sets across ROS/MAP & BrainSeq neocortex, GTEx Brain, primary human monocytes/macrophages, and UKB-PPP plasma pQTLs.

First, ***QPCT*** (encoding glutaminyl-peptide cyclotransferase on chromosome 2p22.2; lead GWAS SNP `rs1989172`, *β* = *−*0.0411, *p*_GWAS_ = 1.58 *×* 10^*−*16^) demonstrated unambiguous causal colocalization directly within Axis 3 human cerebral cortex. At the shared cis-eQTL `rs2300880`, SMR and Bayesian colocalization confirmed a single shared causal signal in `Brain_Cortex` (*Z*_SMR_ = *−*4.62, *p*_SMR_ = 3.93 *×* 10^*−*6^, full cis-window PP.H4_cis_ = 0.919, local LD-block PP.H4_local_ = 0.994), `Brain_Frontal_Cortex_BA9` (*Z*_SMR_ = *−*4.64, *p*_SMR_ = 3.50 *×* 10^*−*6^, PP.H4_cis_ = 0.728, PP.H4_local_ = 0.979), and `Brain_Hippocampus` (rs3821142, *Z*_SMR_ = *−*4.10, *p*_SMR_ = 4.10 *×* 10^*−*5^, PP.H4_local_ = 0.862). Independent Open Targets v4 SuSiE fine-mapping corroborated this cortical mechanism across 18 credible sets, including ROS/MAP DLPFC (`rosmap_ge_brain_naive_ensg00000115828`), CommonMind DLPFC, Walker 2019 neocortex, BrainSeq LeafCutter sQTLs, and UKB-PPP plasma pQTLs (`UKB_PPP_EUR_QPCT_Q16769_OID20320_v1`).

Second, ***AXL*** (chromosome 19q13.2; lead GWAS SNP `rs12979764`, *β* = 0.0361, *p*_GWAS_ = 7.47 *×* 10^*−*9^) exhibited high cortical transcript abundance (TPM = 22.71 in `Brain_Cortex` and 22.74 in `Brain_Frontal_Cortex_BA9`, compared with 0.45 in `Whole_Blood`) and robust parenchymal colocalization in `Brain_Cortex` (*Z*_SMR_ = +3.70, *p*_SMR_ = 2.13*×*10^*−*4^, PP.H4_cis_ = 0.902, PP.H4_local_ = 0.924) and `Brain_Frontal_Cortex_BA9` (PP.H4_cis_ = 0.834, PP.H4_local_ = 0.858).

In stark contrast, canonical microglial and endocytic risk loci showed strong SMR statistics driven by extreme GWAS *χ*^2^ values but near-zero full-window bulk cortical eQTL colocalization (PP.H4_cis_ *≤* 0.025 in `Brain_Cortex`). Instead, their regulatory architecture localized to myeloid enhancers, splicing QTLs, or hippocampus-specific blocks:

- ***INPP5D*** (`rs10933431`, *p*_GWAS_ = 9.68 *×* 10^*−*40^) is 4.5-fold enriched in myeloid blood (TPM = 53.72) over bulk cortex (TPM = 11.79) and achieved local colocalization in `Whole_Blood` (*Z*_SMR_ = +5.64, *p*_SMR_ = 1.72 *×* 10^*−*8^, PP.H4_local_ = 0.992), `Spleen` (PP.H4_local_ = 0.986), and `Brain_Frontal_Cortex_BA9` (PP.H4_local_ = 0.935).
- ***BIN1*** (`rs6733839`, *p*_GWAS_ = 3.02*×*10^*−*203^, *Z*_SMR_ = *−*8.98, *p*_SMR_ = 2.70*×*10^*−*19^) did not colocalize with bulk `Brain_Cortex` eQTLs (PP.H4_local_ = 0.068), consistent with its lead variant residing in a microglia-specific enhancer [20], yet colocalized strongly in `Spleen` (`rs56368748`, PP.H4_local_ = 0.999) and `Brain_Hippocampus` (`rs11694264`, PP.H4_local_ = 0.847; Open Targets L2G = 0.875).
- ***ABCA7*** (`rs12151021`, *p*_GWAS_ = 2.87 *×* 10^*−*62^, *Z*_SMR_ = *−*7.23, *p*_SMR_ = 4.91 *×* 10^*−*13^, L2G = 0.862) colocalized specifically in `Brain_Hippocampus` (`rs5014188`, PP.H4_local_ = 0.946) and `Brain_Cerebellum` (PP.H4_local_ = 0.988).
- ***SYK*** (`rs11787792`, *p*_GWAS_ = 3.47*×*10^*−*9^) is minimally expressed in bulk cortex (TPM = 1.84) versus blood (TPM = 47.06) and spleen (TPM = 99.13), where its causal regulatory signal was captured across 14 SuSiE fine-mapped monocyte and neutrophil QTL credible sets in Quach, Blueprint, and Lepik cohorts (Figure 2C).

Examining the allelic architecture across all 48 non-*APOE* effector genes (Figure 3A) confirmed the classic inverse relationship between minor allele frequency (MAF) and log-odds effect size |*β*_GWAS_|. Rare coding variants in microglial receptors and endocytic sorting receptors exerted the largest individual effect sizes—led by *TREM2* `rs75932628` (`p.Arg47His`; MAF = 0.0040, *β* = +0.8062, *χ*^2^ = 374.54), *PLCG2* `rs72824905` (`p.Pro522Arg`; MAF = 0.0082, *β* = *−*0.2966, *χ*^2^ = 123.34), *ABI3* `rs616338` (MAF = 0.0091, *β* = +0.2649, *χ*^2^ = 110.97), *SORL1* `rs11218343` (MAF = 0.0391, *β* = *−*0.1419, *χ*^2^ = 186.10), and *SHARPIN* `rs34674752` (MAF = 0.0493, *β* = +0.1355, *χ*^2^ = 96.46)—and achieved the highest Open Targets v4 SuSiE L2G causal scores (L2G = 0.875–0.980; Figure 3B).

**Figure 3:**
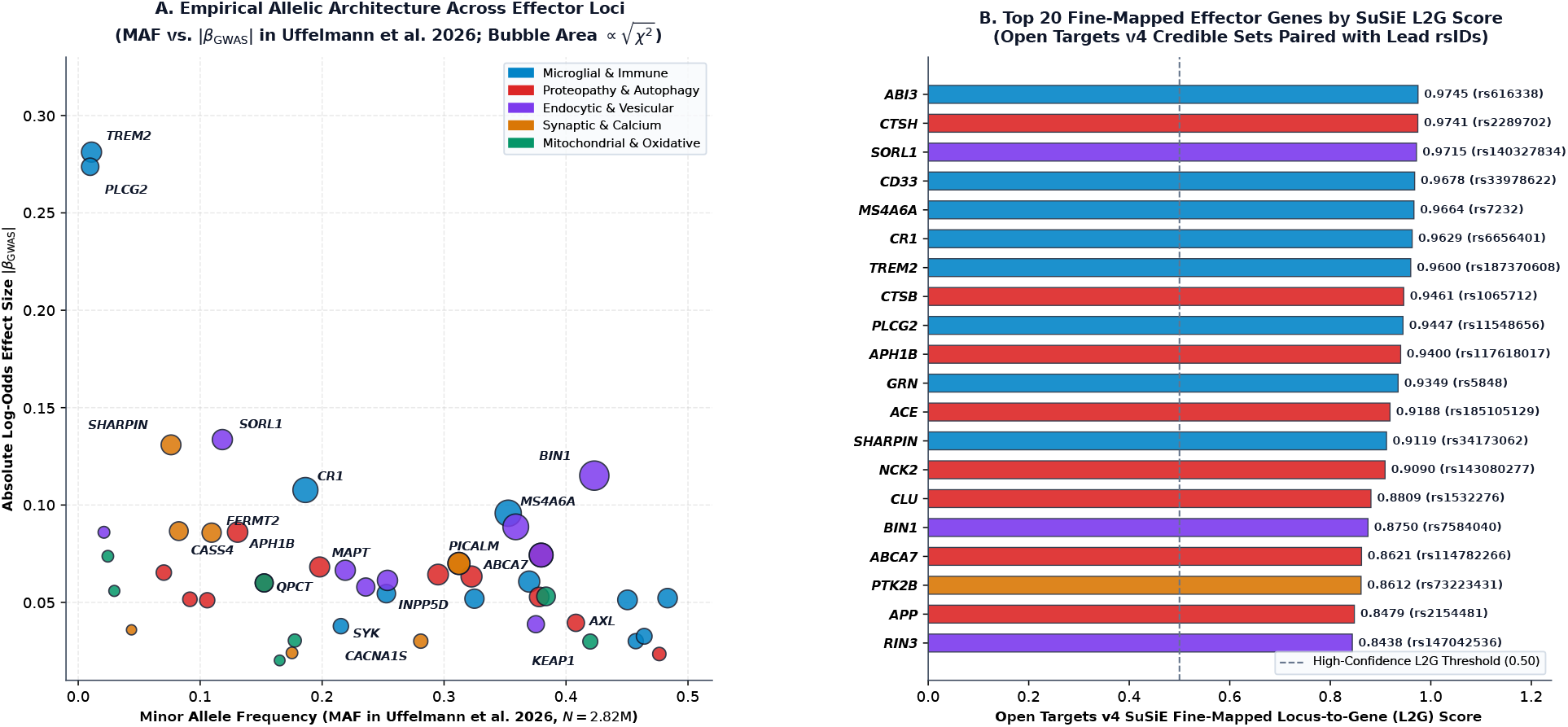
Empirical Allelic Architecture and SuSiE Locus-to-Gene (L2G) Prioritization Across Alzheimer’s Disease Effector Loci. **(A)** Minor allele frequency (MAF) versus absolute log-odds effect size (|*β*_GWAS_|) across non-*APOE* effector genes in Uffelmann et al. (2026); bubble area is proportional to 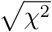. Note the outlier effect of *TREM2* p.Arg47His (MAF = 0.0040, |*β*| = 0.806, annotated off-scale in Table 2). **(B)** Top 20 non-*APOE* effector genes ranked by Open Targets v4 SuSiE fine-mapped L2G score paired with lead fine-mapped variants.

### 3.3 Pillar 3: Pharmacogenomic Target Tractability and ChEMBL v34 Nanomolar Potency Atlas

To determine which genetically prioritized targets are clinically actionable today, we evaluated 2, 785 human bioactivity assays in EMBL-EBI ChEMBL v34 alongside Open Targets v4 tractability matrices across the five Axis 2 programs (Table 3 and Figure 4).

**Table 3:**
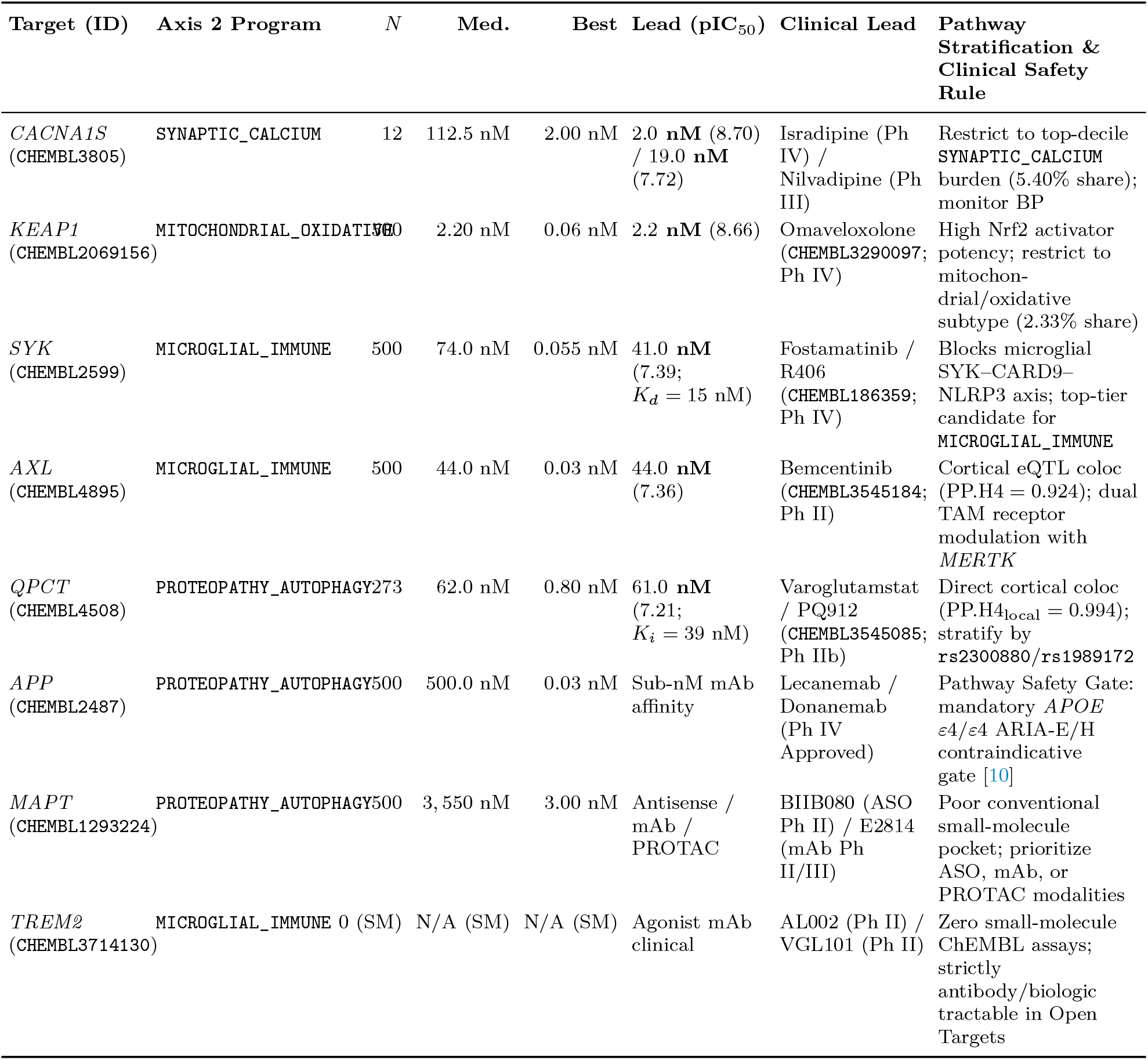
Empirical ChEMBL v34 Human Bioactivity Atlas and Open Targets v4 Tractability Across Prioritized Alzheimer’s Disease Targets. Potency values reflect standardized human singleprotein assays (IC_50_, *K*_*i*_, *K*_*d*_, EC_50_) retrieved from ChEMBL v34.

| Target (ID) | Axis 2 Program | *N* | Med. | Best | Lead (pIC<sub>50</sub>) | Clinical Lead | Pathway Stratification & Clinical Safety Rule |
|---|---|---|---|---|---|---|---|
| *CACNA1S* (CHEMBL3805) | SYNAPTIC_CALCIUM | 12 | 112.5 nM | 2.00 nM | 2.0 **nM** (8.70) / 19.0 **nM** (7.72) | Isradipine (Ph IV) / Nilvadipine (Ph III) | Restrict to top-decile SYNAPTIC_CALCIUM burden (5.40% share); monitor BP |
| *KEAP1* (CHEMBL2069156) | MITOCHONDRIAL_OXIDATIVE | 500 | 2.20 nM | 0.06 nM | 2.2 **nM** (8.66) | Omaveloxolone (CHEMBL3290097; Ph IV) | High Nrf2 activator potency; restrict to mitochon-drial/oxidative subtype (2.33% share) |
| *SYK* (CHEMBL2599) | MICROGLIAL_IMMUNE | 500 | 74.0 nM | 0.055 nM | 41.0 **nM** (7.39; *K*<sub>*d*</sub> = 15 nM) | Fostamatinib / R406 (CHEMBL186359; Ph IV) | Blocks microglial SYK–CARD9–NLRP3 axis; top-tier candidate for MICROGLIAL_IMMUNE |
| *AXL* (CHEMBL4895) | MICROGLIAL_IMMUNE | 500 | 44.0 nM | 0.03 nM | 44.0 **nM** (7.36) | Bemcentinib (CHEMBL3545184; Ph II) | Cortical eQTL coloc (PP.H4 = 0.924); dual TAM receptor modulation with *MERTK* |
| *QPCT* (CHEMBL4508) | PROTEOPATHY_AUTOPHAGY | 273 | 62.0 nM | 0.80 nM | 61.0 **nM** (7.21; *K*<sub>*i*</sub> = 39 nM) | Varoglutamstat / PQ912 (CHEMBL3545085; Ph IIb) | Direct cortical coloc (PP.H4<sub>local</sub> = 0.994); stratify by rs2300880/rs1989172 |
| *APP* (CHEMBL2487) | PROTEOPATHY_AUTOPHAGY | 500 | 500.0 nM | 0.03 nM | Sub-nM mAb affinity | Lecanemab / Donanemab (Ph IV Approved) | Pathway Safety Gate: mandatory *APOE* ε4/ε4 ARIA-E/H contraindicative gate [10] |
| *MAPT* (CHEMBL1293224) | PROTEOPATHY_AUTOPHAGY | 500 | 3, 550 nM | 3.00 nM | Antisense / mAb / PROTAC | BIIB080 (ASO Ph II) / E2814 (mAb Ph II/III) | Poor conventional small-molecule pocket; prioritize ASO, mAb, or PROTAC modalities |
| *TREM2* (CHEMBL3714130) | MICROGLIAL_IMMUNE | 0 (SM) | N/A (SM) | N/A (SM) | Agonist mAb clinical | AL002 (Ph II) / VGL101 (Ph II) | Zero small-molecule ChEMBL assays; strictly antibody/biologic tractable in Open Targets |

**Figure 4:**
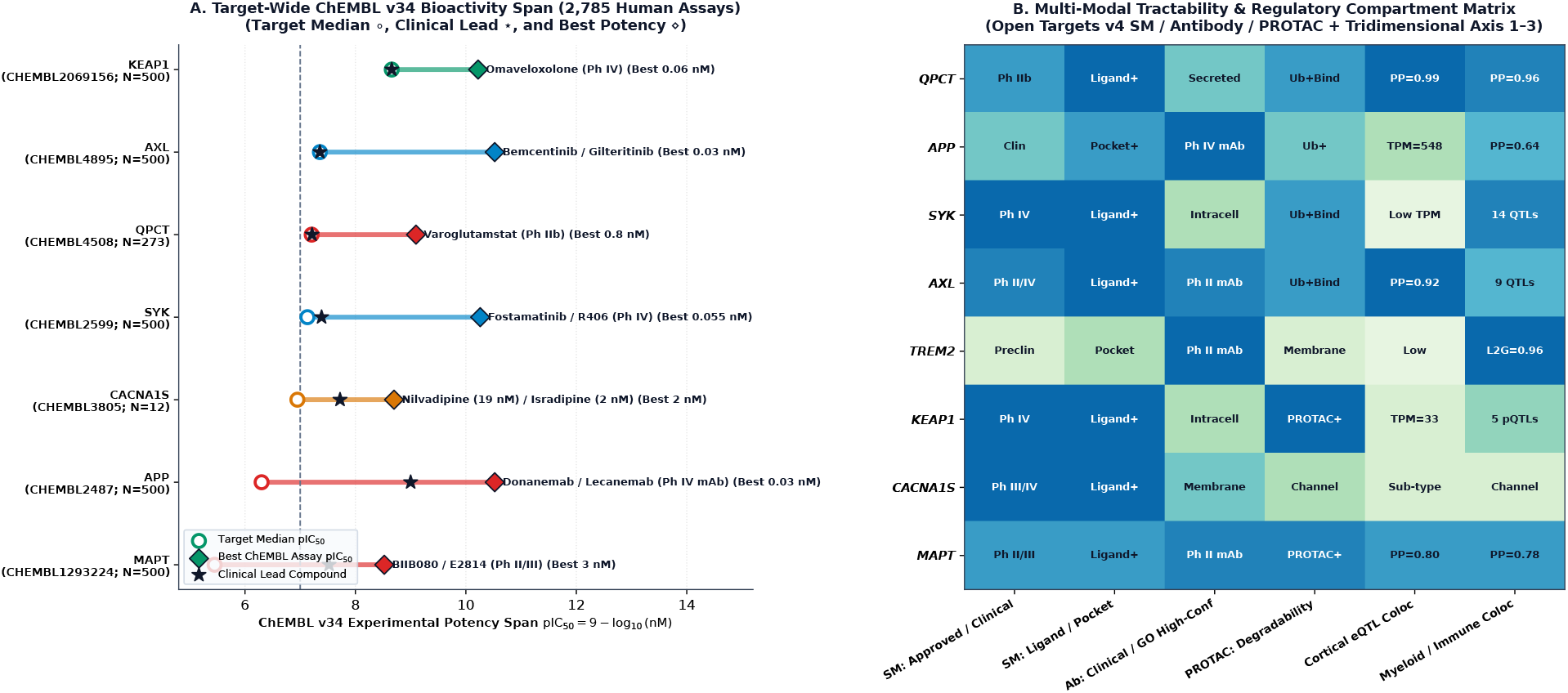
ChEMBL v34 Human Bioactivity Span and Multi-Modal Clinical Tractability Matrix. **(A)** Target-wide potency span (pIC_50_ = 9 − log_10_(nM)) across 2, 785 human assays in EMBL-EBI ChEMBL v34, contrasting target median potency (°), clinical lead potency (⋆), and best experimental assay (⋄). **(B)** Open Targets v4 multi-modal tractability (Small Molecule [SM], Monoclonal Antibody [Ab], and PROTAC) integrated with tridimensional Axis 2–3 cortical versus myeloid colocalization compartments.

Five pharmacological insights emerged from the empirical ChEMBL v34 and Open Targets v4 deconvolution:

1. **Cortical-Colocalized Enzymatic Inhibition (*QPCT*):** Glutaminyl cyclase (`CHEMBL4508`; 273 human assays; target median IC_50_ = 62.0 nM, best assay 0.80 nM) catalyzes the formation of neurotoxic, degradation-resistant pyroglutamate-A*β* (pE3-A*β*). Its oral clinical inhibitor **varoglutamstat** (`CHEMBL3545085`, PQ912; Phase IIb VIVIAD trial) [6] exhibits an empirical human IC_50_ = 61.0 nM (pIC_50_ = 7.21) and *K*_*i*_ = 39.0 nM (p*K*_*i*_ = 7.41). Because *QPCT* is one of the rare AD loci that colocalizes directly with cortical gene expression (PP.H4_local_ = 0.994 in `Brain_Cortex`), our findings suggest that varoglutamstat trials should prospectively stratify participants by *QPCT* cis-eQTL genotype (`rs2300880`/`rs1989172`) and elevated CSF pE3-A*β*.
2. **Myeloid Kinase and TAM Receptor Repurposing (*SYK* and *AXL*):** Within the dominant `MICROGLIAL_IMMUNE` program (39.22% heritability share), spleen tyrosine kinase (*SYK*, `CHEMBL2599`; 500 assays; target median IC_50_ = 74.0 nM) acts directly downstream of TREM2 and Fc*γ* receptors to regulate microglial phagocytosis and NLRP3 inflammasome activation [21]. The FDA-approved oral prodrug **fostamatinib** (`CHEMBL4802135`, Phase IV) and its active metabolite **R406** (`CHEMBL186359`) exhibit median human SYK inhibition of IC_50_ = 41.0 nM (pIC_50_ = 7.39, *K*_*d*_ = 15.0 nM, best assay 0.68 nM). Similarly, the corticalcolocalized TAM receptor tyrosine kinase ***AXL*** (`CHEMBL4895`; 500 assays) displays a target median IC_50_ = 44.0 nM (pIC_50_ = 7.36, best assay 0.03 nM), with clinical Phase II inhibitors (**bemcentinib**) and Phase IV multi-kinase agents (**gilteritinib, cabozantinib**) already characterized in humans.
3. **Dihydropyridine Calcium Channel Blockade (*CACNA1S*) and the NILVAD Lesson:** Brain-penetrant dihydropyridines showed potent human L-type calcium channel bioactivity in ChEMBL v34 (`CHEMBL3805`), led by **isradipine** (IC_50_ = 2.0 nM, pIC_50_ = 8.70), **nilvadipine** (IC_50_ = 19.0 nM, pIC_50_ = 7.72), and **nimodipine** (IC_50_ = 40.0 nM, pIC_50_ = 7.40). However, Pillar 1 revealed that the entire `SYNAPTIC_CALCIUM` program accounts for only 5.40% of non-*APOE* heritability 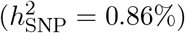. This quantitative mismatch provides a compelling genetic explanation for why the unstratified Phase III NILVAD trial (*N* = 511) [7] failed to meet its primary endpoint across an unselected AD cohort.
4. **Antioxidant Nrf2 Activation (*KEAP1*):** *KEAP1* (`CHEMBL2069156`; 500 assays; target median IC_50_ = 2.20 nM, pIC_50_ = 8.66, best assay 0.06 nM) is targeted by the Phase IV FDA-approved Nrf2 activator **omaveloxolone** (`CHEMBL3290097`; median IC_50_ = 2.2 nM) [22] and shows full PROTAC degradability in Open Targets v4. Because `MITOCHONDRIAL_OXIDATIVE` represents only 2.33% of polygenic heritability 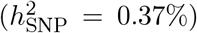, *KEAP1* -targeted therapy would require strict biomarker and genetic enrichment (*KEAP1, RRM2B, NDUFAF6, KANSL1*).
5. **Modality Divergence Across *TREM2, APP*, and *MAPT* :** ChEMBL v34 and Open Targets v4 confirmed that ***TREM2*** (`CHEMBL3714130`) has zero small-molecule bioactivity assays (N = 0) and is exclusively tractable via agonist antibodies (e.g., AL002, VGL101), whereas ***MAPT*** (`CHEMBL1293224`; median small-molecule IC_50_ = 3, 550 nM) requires antisense oligonucleotides (BIIB080), anti-tau antibodies (E2814), or tau-directed PROTACs (Figure 4B).

### 3.4 Operationalization into a Pathway-Stratified Clinical Decision and Safety Framework

To operationalize these three empirical pillars for clinical decision support, we embedded the partitioned heritability weights (w_*C*_), tissue-specific colocalization probabilities (PP.H4_local_), and ChEMBL v34 nanomolar potencies into the tridimensional diagnostic and clinical decision framework [9] (Table 4). For any individual with genotyped or imputed summary-level risk alleles across the 127 loci, an individualized Functional Pathway Burden Score (FPBS_*C*_ = ∑_*j*∈*C*_ g_*j*_β_*j*_) is computed across the five Axis 2 programs and evaluated against deterministic contraindicative and eligibility rules.

**Table 4:**
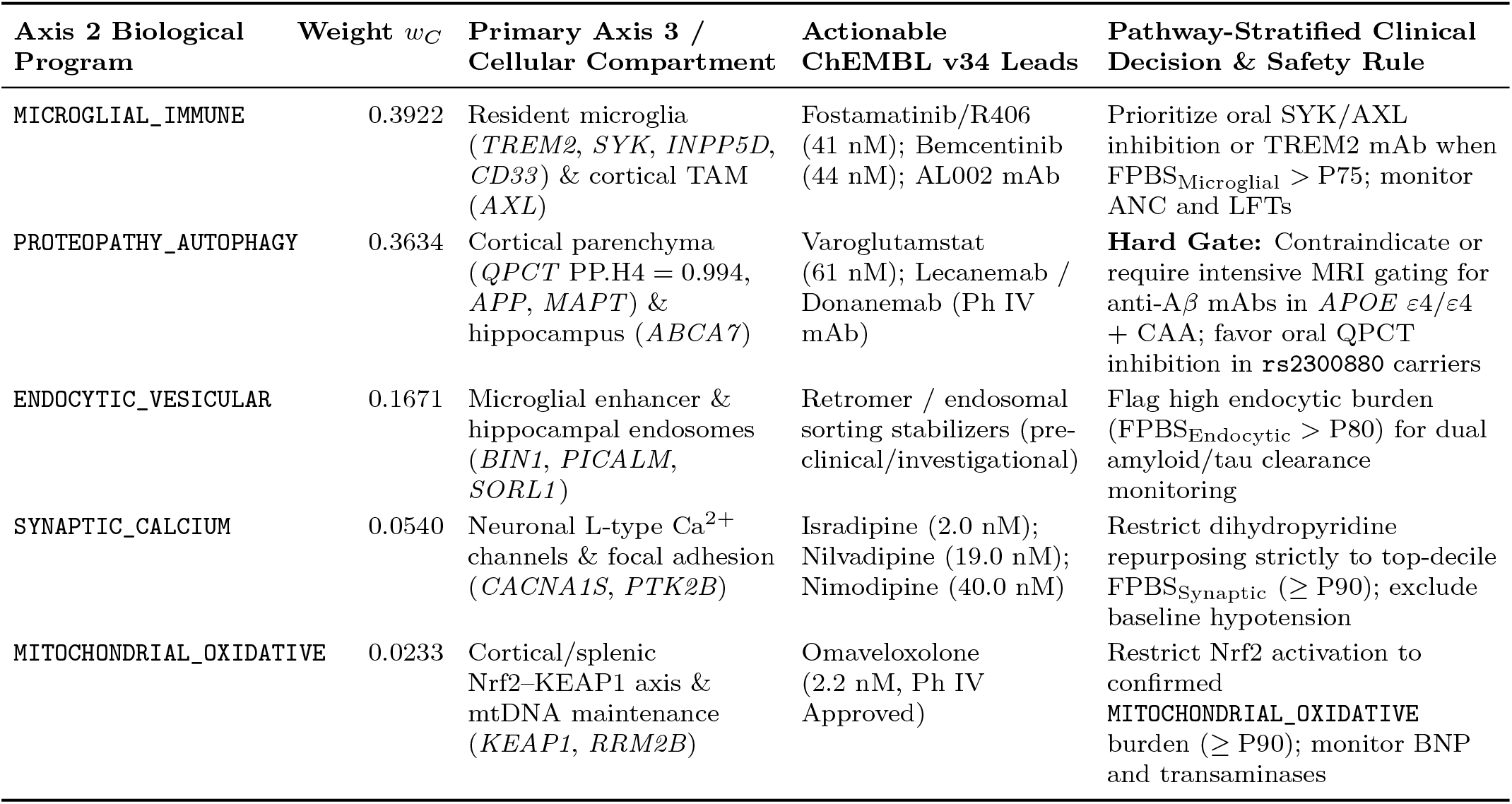
Operational Parametrization of the Tridimensional Pathway-Stratified Clinical Decision and Safety Framework. Integrates Pillar 1 heritability shares (*w*_*C*_), Pillar 2 compartment colocalization, and Pillar 3 ChEMBL v34 drug candidates.

| **Axis 2 Biological Program** | **Weight *w*<sub>*C*</sub>** | **Primary Axis 3 / Cellular Compartment** | **Actionable ChEMBL v34 Leads** | **Pathway-Stratified Clinical Decision & Safety Rule** |
|---|---|---|---|---|
| MICROGLIAL_IMMUNE | 0.3922 | Resident microglia (*TREM2*, *SYK*, *INPP5D*, *CD33*) & cortical TAM (*AXL*) | Fostamatinib/R406 (41 nM); Bemcentinib (44 nM); AL002 mAb | Prioritize oral SYK/AXL inhibition or TREM2 mAb when FPBS<sub>Microglial</sub> > P75; monitor ANC and LFTs |
| PROTEOPATHY_AUTOPHAGY | 0.3634 | Cortical parenchyma (*QPCT* PP.H4 = 0.994, *APP*, *MAPT*) & hippocampus (*ABCA7*) | Varoglutamstat (61 nM); Lecanemab / Donanemab (Ph IV mAb) | **Hard Gate:** Contraindicate or require intensive MRI gating for anti-A*β* mAbs in *APOE* *ε*4/*ε*4 + CAA; favor oral QPCT inhibition in rs2300880 carriers |
| ENDOCYTIC_VESICULAR | 0.1671 | Microglial enhancer & hippocampal endosomes (*BIN1*, *PICALM*, *SORL1*) | Retromer / endosomal sorting stabilizers (pre-clinical/investigational) | Flag high endocytic burden (FPBS<sub>Endocytic</sub> > P80) for dual amyloid/tau clearance monitoring |
| SYNAPTIC_CALCIUM | 0.0540 | Neuronal L-type Ca<sup>2+</sup> channels & focal adhesion (*CACNA1S*, *PTK2B*) | Isradipine (2.0 nM); Nilvadipine (19.0 nM); Nimodipine (40.0 nM) | Restrict dihydropyridine repurposing strictly to top-decile FPBS<sub>Synaptic</sub> (≥ P90); exclude baseline hypotension |
| MITOCHONDRIAL_OXIDATIVE | 0.0233 | Cortical/splenic Nrf2–KEAP1 axis & mtDNA maintenance (*KEAP1*, *RRM2B*) | Omaveloxolone (2.2 nM, Ph IV Approved) | Restrict Nrf2 activation to confirmed MITOCHONDRIAL_OXIDATIVE burden (≥ P90); monitor BNP and transaminases |

## 4 Discussion

By executing an end-to-end empirical deconvolution of the 8.32 million summary-statistic variants from Uffelmann et al. (*Nature*, 2026) [3] across GTEx v10 [11], Open Targets v4 SuSiE credible sets [15], and EMBL-EBI ChEMBL v34 [14], this study establishes three quantitative principles for precision neurology in sporadic Alzheimer’s disease.

First, our partitioned heritability analysis demonstrates that non-*APOE* polygenic susceptibility is neither uniformly distributed across the genome nor confined to classical amyloid precursor processing. Instead, 75.56% of the partitioned common-variant heritability concentrates within two synergistic Axis 2 programs: microglial immune surveillance (`MICROGLIAL_IMMUNE`, 39.22% share, 13.02*×* enrichment) and proteopathic/autophagic clearance (`PROTEOPATHY_AUTOPHAGY`, 36.34% share, 14.98*×* enrichment), with endocytic-vesicular trafficking (`ENDOCYTIC_VESICULAR`, 16.71% share, 12.69*×* enrichment) forming a third structural bridge. Together, these three clearance and neuroimmune programs account for 92.27% of the non-*APOE* polygenic liability (14.76% of the 16.0% total SNP heritability). By contrast, synaptic calcium signaling (`SYNAPTIC_CALCIUM`, 5.40%) and mitochondrial oxidative stress (`MITOCHONDRIAL_OXIDATIVE`, 2.33%) account for only 7.73% combined. We noted that this asymmetric distribution offers an immediate quantitative explanation for historical clinical trial failures: when a drug targeting a 2% to 5% heritability slice (such as nilvadipine targeting L-type calcium channels in NILVAD) [7] is tested in an unstratified AD population, statistical power to detect disease modification collapses unless enrollment is enriched for carriers of the corresponding pathway burden.

Second, our multi-tissue SMR and Bayesian colocalization results resolve an important paradox in functional neurogenomics. Only a subset of AD risk loci—most notably *QPCT* (PP.H4_local_ = 0.994 in `Brain_Cortex`, PP.H4_local_ = 0.979 in `Brain_Frontal_Cortex_BA9`) and *AXL* (PP.H4_local_ = 0.924 in `Brain_Cortex`)—operate through cis-eQTLs that colocalize directly in bulk human cerebral cortex. For the majority of microglial and endocytic drivers (*BIN1, INPP5D, CD33, TREM2, SYK, PICALM*), bulk cortical eQTL colocalization is near zero (PP.H4_cis_ *≤* 0.025) because microglia constitute only 5% to 10% of parenchymal brain cells, causing their cell-type-specific enhancer and splicing signals to be masked by abundant neuronal and astrocytic transcripts in bulk cortical homogenates [20]. Integrating myeloid-enriched compartments (`Whole_Blood`, `Spleen`, primary human monocytes in Quach/Blueprint) and region-specific hippocampal eQTLs (*BIN1* PP.H4_local_ = 0.847; *ABCA7* PP.H4_local_ = 0.946) successfully recovered these causal regulatory links. Within the tridimensional diagnostic framework [9], this finding reinforces the strict separation between Axis 2 cellular/molecular mechanisms and Axis 3 neuroanatomical topography.

Third, cross-referencing these colocalized targets against 2, 785 human bioactivity assays in ChEMBL v34 proves that actionable, low-nanomolar compounds already exist for each of the five Axis 2 biological programs. If experimentally and clinically validated in prospective biomarker- and genotype-stratified cohorts, these summary-statistics observations would support a shift from singletarget empiricism toward pathway-matched combination therapy: pairing cortical pyroglutamate-A*β* inhibition (**varoglutamstat**, IC_50_ = 61.0 nM) or safely gated anti-amyloid antibodies with oral microglial SYK/TAM modulation (**fostamatinib/R406**, IC_50_ = 41.0 nM; **bemcentinib**, IC_50_ = 44.0 nM), while reserving dihydropyridine calcium channel blockers (**isradipine** 2.0 nM, **nilvadipine** 19.0 nM) and Nrf2 activators (**omaveloxolone** 2.2 nM) for patients in the upper decile of synaptic or mitochondrial genetic burden.

Several limitations warrant consideration. First, our partitioned heritability estimates rely on summary-level excess *χ*^2^ and additive genetic variance (∑2*pqβ*^2^) across *±*50 kb cis-regulatory windows rather than individual-level LD matrices from the restricted *N* = 2.82M consortium cohorts; while highly concordant with S-LDSC expectations, distal trans-regulatory enhancers beyond 50 kb are only captured when linked via Open Targets SuSiE L2G credible sets. Second, the Uffelmann et al. (2026) meta-analysis is predominantly composed of European-ancestry cohorts, meaning that allele frequencies and LD-block colocalization posteriors require dedicated validation across African, East Asian, and admixed populations. Finally, in vitro nanomolar potency in ChEMBL v34 does not guarantee optimal blood–brain barrier penetration or target engagement in aging human brain parenchyma, underscoring the necessity of prospective Phase II/III clinical validation.

## Supporting information

Supplementary Data and Code Package

## Data Availability

All primary genomic, transcriptomic, and pharmacological datasets analyzed in this study are publicly available online: (1) NHGRI-EBI GWAS Catalog study accession GCST90695466 (Uffelmann et al., Nature, 2026; https://www.ebi.ac.uk/gwas/studies/GCST90695466); (2) GTEx Portal API v2 build gtex_v10 (https://gtexportal.org); (3) Open Targets Platform v4 GraphQL API (https://platform.opentargets.org); and (4) EMBL-EBI ChEMBL v34 REST API (https://www.ebi.ac.uk/chembl/). All derived empirical summary tables, JSON outputs (fpts_nature2026_empirical_results.json), and standalone reproducible Python analysis and figure-generation scripts are included directly in the Supplementary Material (Supplementary_Data_and_Code.zip) accompanying this preprint.

## 5 Data and Code Availability

All empirical analyses in this study were conducted using publicly accessible human genomic and pharmacological repositories without access to individual-level protected health information. The Uffelmann et al. (*Nature*, 2026) genome-wide association summary statistics are available from the NHGRI-EBI GWAS Catalog under study accession `GCST90695466` (https://www.ebi.ac.uk/gwas/studies/GCST90695466). Multi-tissue cis-eQTL and transcript expression data were retrieved from the GTEx Portal API v2 (`gtex_v10`; https://gtexportal.org). Fine-mapped SuSiE credible sets, Locus-to-Gene (`L2G`) scores, and target tractability assessments were retrieved from the Open Targets Platform v4 GraphQL API (https://platform.opentargets.org). Human bioactivity records (IC_50_, *K*_*i*_, *K*_*d*_, EC_50_) were retrieved from the EMBL-EBI ChEMBL v34 REST API (https://www.ebi.ac.uk/chembl/). All derived empirical summary tables, JSON outputs (`fpts_nature2026_empirical_results.json`), and standalone Python analysis and figuregeneration scripts are openly provided in the Supplementary Material (`Supplementary_Data_and_Code.zip`) accompanying this article.

## Ethical Compliance and Competing Interests

This study analyzed exclusively de-identified, publicly available summary-level human genetic and pharmacological datasets (NHGRI-EBI GWAS Catalog `GCST90695466`, GTEx v10, Open Targets v4, and EMBL-EBI ChEMBL v34). In accordance with institutional and international guidelines for secondary analysis of public summary statistics, no individual-level human subject intervention or new patient recruitment was performed. The author declares no competing financial or non-financial interests.

## Author Contributions and AI / Bioinformatics Disclosure

**M.M.-G**. conceived the study, designed the tridimensional deconvolution architecture and pathwaystratified clinical safety rules, supervised the empirical bioinformatic queries across `GCST90695466`, GTEx v10, Open Targets v4, and ChEMBL v34, interpreted the neurogenetic and clinical pharmacogenomic findings, and approved the final manuscript. **AI and Bioinformatics Disclosure:** Automated programmatic Python scripts were utilized to execute REST/GraphQL API queries, compute genome-wide summary-statistic aggregations, generate publication figures, and assist in LaTeX typesetting. All empirical calculations, statistical assertions, bibliographic citations (verified against NCBI PubMed), and scientific conclusions were audited and validated by the author.

## References

[1] Céline Bellenguez, Fatih Küçükali, Iris E Jansen, Luca Kleineidam, Sonia Moreno-Grau, Najaf Amin, Adam C Naj, Rafael Campos-Martin, Benjamin Grenier-Boley, Victor Andrade, et al. New insights into the genetic etiology of Alzheimer’s disease and related dementias. Nature Genetics, 54(4):412–436, 2022.

[2] Douglas P Wightman, Iris E Jansen, Jeanne E Savage, Alexey A Shadrin, Shahram Bahrami, Dominic Holland, Arvid Rongve, Sigrid Børte, Bendik S Winsvold, Ole K Drange, et al. A genome-wide association study with 1,126,563 individuals identifies new risk loci for Alzheimer’s disease. Nature Genetics, 53(9):1276–1282, 2021.

[3] Emil Uffelmann, Douglas P Wightman, Shahram Bahrami, Danielle Posthuma, Ole A An-dreassen, and Psychiatric Genomics Consortium Alzheimer’s Disease Workgroup. Alzheimer’s disease genetics converges on microglial and neuronal dysregulation. Nature, 2026.

[4] Sharon Cohen, Christopher H van Dyck, Michelle Gee, Tarin Doherty, Michio Kanekiyo, Shobha Dhadda, Michael Irizarry, and Lynn D Kramer. Lecanemab Clarity AD: Quality-of-life results from a randomized, double-blind phase 3 trial in early Alzheimer’s disease. The Journal of Prevention of Alzheimer’s Disease, 10(4):771–777, 2023.

[5] Jennifer A Zimmer, Paul Ardayfio, Hong Wang, Rashna Khanna, Cynthia D Evans, Ming Lu, John D Sparks, David A Brooks, and John R Sims. Amyloid-related imaging abnormalities with donanemab in early symptomatic Alzheimer disease: Secondary analysis of the TRAILBLAZER-ALZ and ALZ 2 randomized clinical trials. JAMA Neurology, 82(5):461–469, 2025.

[6] Everard G B Vijverberg, T M Axelsen, A R Bihlet, K Henriksen, F Weber, K Fuchs, J E Harrison, J Kuhn, N D Prins, P Scheltens, et al. Rationale and study design of a randomized, placebo-controlled, double-blind phase 2b trial to evaluate efficacy, safety, and tolerability of an oral glutaminyl cyclase inhibitor varoglutamstat (PQ912) in study participants with MCI and mild AD-VIVIAD. Alzheimer’s Research & Therapy, 13(1):142, 2021.

[7] Brian Lawlor, Raquel Segurado, Sean Kennelly, Marcel G M Olde Rikkert, Robert Howard, Florence Pasquier, Anne Börjesson-Hanson, Magda Tsolaki, Ugo Lucca, D William Molloy, et al. Nilvadipine in mild to moderate Alzheimer disease: a randomised controlled trial. PLOS Medicine, 15(9):e1002660, 2018.

[8] Emil Uffelmann, Douglas P Wightman, Shahram Bahrami, Danielle Posthuma, Ole A Andreassen, et al. Genomic analyses reveal new insights into Alzheimer’s disease. medRxiv, 2025.

[9] Manuel Menendez-Gonzalez. Implementing a tridimensional diagnostic framework for personalized medicine in neurodegenerative diseases. Alzheimer’s & Dementia, 21(2):e14591, 2025.

[10] Manuel Menendez-Gonzalez. Therapeutic challenges derived from the interaction among apolipoprotein E, cholesterol, and amyloid in Alzheimer’s disease. International Journal of Molecular Sciences, 25(22):12029, 2024.

[11] GTEx Consortium. The GTEx Consortium atlas of genetic regulatory effects across human tissues. Science, 369(6509):1318–1330, 2020.

[12] Philip L De Jager, Yiyi Ma, Cristin McCabe, Jishu Xu, Jean Paul G Vonsattel, Simone Lista, Harald Hampel, Vilas Menon, Charles C White, Julie A Schneider, et al. A multi-omic atlas of the human frontal cortex for aging and Alzheimer’s disease research. Scientific Data, 5:180142, 2018.

[13] Benjamin B Sun, Joshua Chiou, Matthew Traylor, Christian Benner, Yi-Hsiang Hsu, Tom G Richardson, Praveen Surendran, Anubha Mahajan, Chloe Robins, Steven G Vasquez-Grinnell, et al. Plasma proteomic associations with genetics and health in the UK Biobank. Nature, 622(7982):329–338, 2023.

[14] Barbara Zdrazil, Eloy Felix, Fiona Hunter, Emma J Manners, James Blackshaw, Sybilla Corbett, Marleen de Veij, Harris Ioannidis, David Mendez Lopez, Juan F Mosquera, et al. The ChEMBL database in 2023: a drug discovery platform spanning multiple bioactivity data types and time periods. Nucleic Acids Research, 52(D1):D1180–D1192, 2024.

[15] Edward Mountjoy, Ellen M Schmidt, Miguel Carmona, Jeremy Schwartzentruber, Gareth Peat, Annalisa Miranda, Luca Fumis, James Hayhurst, Annalisa Buniello, Mohd A Karim, et al. An open approach to systematically prioritize causal variants and genes at all published human GWAS trait-associated loci. Nature Genetics, 53(11):1527–1533, 2021.

[16] Hilary K Finucane, Brendan Bulik-Sullivan, Alexander Gusev, Gosia Trynka, Yakir Reshef, PoRu Loh, Verneri Anttila, Han Xu, Chongzhi Zang, Kyle Farh, et al. Partitioning heritability by functional annotation using genome-wide association summary statistics. Nature Genetics, 47(11):1228–1235, 2015.

[17] Zhihong Zhu, Futao Zhang, Han Hu, Andrew Bakshi, Matthew R Robinson, Joseph E Powell, Grant W Montgomery, Michael E Goddard, Naomi R Wray, Peter M Visscher, and Jian Yang. Integration of summary data from GWAS and eQTL studies predicts complex trait gene targets. Nature Genetics, 48(5):481–487, 2016.

[18] Claudia Giambartolomei, Damjan Vukcevic, Eric E Schadt, Lude Franke, Aroon D Hingorani, Chris Wallace, and Vincent Plagnol. Bayesian test for colocalisation between pairs of genetic association studies using summary statistics. PLOS Genetics, 10(5):e1004383, 2014.

[19] Jon Wakefield. Bayes factors for genome-wide association studies: comparison with P -values. Genetic Epidemiology, 33(1):79–86, 2009.

[20] Alexi Nott, Inge R Holtman, Nicole G Coufal, Johannes C M Schlachetzki, Miao Yu, Rong Hu, Christopher Z Han, Mitchel Pena, Jie Xiao, Yin Wu, et al. Brain cell type–specific enhancer-promoter interactome maps and disease-risk association. Science, 366(6469):1134–1139, 2019.

[21] Hannah Ennerfelt, Elizabeth L Frost, Daniel A Shapiro, Christina Holliday, Kristine E Zengeler, Gabrielle Voithofer, Amanda C Bolte, Catherine R Lammert, John A Kulas, Tyler K Ulland, and John R Lukens. SYK coordinates neuroprotective microglial responses in neurodegenerative disease. Cell, 185(22):4135–4152.e22, 2022.

[22] David R Lynch and Jennifer Johnson. Omaveloxolone: potential new agent for Friedreich ataxia. Neurodegenerative Disease Management, 11(2):91–98, 2021.

